# A Phylogenomic Analysis of HIV Transmission Pattern among High Risk Groups of North-West India

**DOI:** 10.1101/2023.02.27.23286499

**Authors:** Chandar Kanta Chauhan, P.V.M. Lakshmi, Phulen Sarma, Vivek Sagar, Aman Sharma, Sunil K.Arora, Rajesh Kumar

## Abstract

**Background:** Molecular techniques can enhance the power of epidemiological investigations for tracing HIV transmission networks. This information could be useful for developing strategies for prevention of HIV transmission. Hence, we carried out to a study on the transmission patterns among newly diagnosed HIV cases among High-Risk Groups (HRGs) of North-West India using phylogenomic methods.

**Methods:** Phylogenomic analysis was carried out among 37 randomly selected samples of recently infected HRGs identified through Recent Infections Testing Algorithm (RITA) using Limiting Antigen Avidity Assay. Amplification of the reverse transcriptase region of *pol* gene (540 base pairs) and sequencing was done. Reference sequences were extracted from HIV Los Alamos database. Sequences aligned by Clustal W and HIV-1 subtype were determined on the basis of phylogenomic analysis of the *pol* sequence. Phylogenetic trees were constructed using the MEGA (version 11.0).

**Results:** The phylogeny clearly depicts that the study isolates RTFSWCHD and RTFSWPB007 cluster with and are related to the Indian reference sequences AY746371 and EU683781 and a Nepalese sequence KX430115.The other study isolates (RTFSWCHD001, RTFSWPB005, RTFSWCHD002, RTFSWPB006, RTFSWHR008, RTFSWHR 009) clustered uniquely among themselves without any interlinking with other references. One study isolate (RTFSWHP004) clustered closely with Zimbabwian isolate AY998351. The phylogeny shows that the study isolate MSMCHD005 clades separately with the Indian references (DQ838761, EU683781and AY746371), but is also very closely related to the references from China (HG421606, JQ658754), Nepal(JN023039) and Myanmar (N223216, JN223183, KC913773). Other study isolates (MSMCHD003, MSMHP007, MSMCHD004, MSMPB001, MSMPB002, and MSMHR006) are highly interrelated among themselves and form a separate unique clade together. The evolutionary tree shows that all the sequences from current study formed a monophyletic lineage, i.e., sequences from India clustered together more than with sequences from any other country. The study sequences showed relatedness only to the Nepal references KX430115 and JN023035. The South African, UK, Norway, China, and Myanmar references are grouped into aseparate clade.

**Conclusion:** Molecular epidemiologic methods were able to reveal transmission networks; hence, phylogenomic methods can be used in HIV Sentinel Surveillance to monitor transmission networks.

## Introduction

Like other sexually transmitted infections Human Immunodeficiency Virus (HIV) infection also transmits through contact imparting a pivotal role to contact tracing in describing the etiology of AIDS[1].At population level, molecular epidemiology tools have been employed to describe closely associated transmission networks linked with sexually transmitted infections as well as with drug resistance. Effectively recognizing highly linked clusters can assist in identifying transmission net works. Tracing these network routes could be very helpful for public health efforts to channelize prevention strategies.

The genetic makeup of HIV turns out to be relatively distinct to each infected individual. Hence, by utilizing data of genetic relatedness of HIV between individuals transmission patterns can be established [2]. And a detailed knowledge of the transmission dynamics of individual epidemics can help the development, execution and continuation of treatment and prevention interventions [3]. Molecular phylogenomic analysis coupled with epidemiological and demographical data can help in providing better insight to understand the dynamics of individual epidemics.

In recent years, HIV phylogenetic tree analysis has gained popularity as a potent tool for studying HIV transmission trends among different populations. Phylogenetic tree, as the name suggests is a tree with leaves, branches, and nodes each of which a metaphor for a specific genetic component ultimately helps in establishing a transmission network within a cluster [4].A viral gene sequence can be sampled to reconstruct phylogenetic tree, which in turn finds application in analyzing course of viral epidemic with time as well as to study past structure of a population. Phylogenetic tree reconstruction has been a favorite approach because it has not only helped epidemiological purposes but has also provided efficient solution in legal cases.

As phylogenomic analyses provides important information about HIV transmission which can be exploited for zeroing on particular locations which are HIV transmission hotspots [4], we studied transmission patterns among newly diagnosed HIV cases from North-West India using phylogenomic methods.

## Methodology

This study was conducted in Anti Retro-viral Therapy (ART) Centres of North-West India (Chandigarh, Amritsar, Jalandhar, Ludhiana and Rohtak)from September 2014 to June 2016.The study protocol was approved by the Institute Ethics Committee, Postgraduate Institute of Medical Education and Research, Chandigarh (HistoPath/14/2925/525; Date: 07/08/2014). Written informed consent was obtained from all the study subjects before enrollment in the study.

### Study population

Only HIV positive individuals registered in last one year, who had not received any antiretroviral therapy were ≥ 18 years of age and whose CD4+ count was ≥200 cells/µl were included in the study.Patients on Directly Observed Treatment, Short Course (DOTS) for Tuberculosis were excluded from the study.

### Collection of the specimens

Study participants were interviewed using semi-structured questionnaire and information about their socio-demographic characteristics, behavioural risk factors treatment history especially about intake of anti-retroviral drugs and CD4 cell counts were collected. From each individual, 6 ml of venous blood was collected in a K_3_EDTA vacutainer tube (Becton Dickinson, USA). Out of the 6ml, 2ml blood was used for CD4 count estimation at respective ART centres. The remaining 4 ml blood was centrifuged at 2500 rpm for 10 minutes at room temperature for obtaining the plasma. The plasma was aspirated using a Pasteur pipette and stored in separate vials at -80°C.

### Sample size

Out of the 64 recently infected HRGs^5^, due to the budget constraints 37 samples were randomly selected (Computer *Random Number Generator* software) for phylogenomic analysis. Of the 37, nine isolates belonged to female sex workers (FSWs), seven were from Men who have sex with Men (MSM) and 21 were from injecting drug users (IDU).

### RNA extraction

Viral RNA was extracted from 560 μl of plasma using the QIAamp Viral RNA Mini kit (Qiagen,Valencia, CA,United States) according to the manufacturer’s instructions. RNA was recovered from the spin columns in a final elution volume of approximately 60 μl.

### PCR amplification and sequencing

The HIV-1 *pol* gene (reverse transcriptase 1–268 codon) was amplified by using nested reverse transcription polymerase chain reaction (RT-PCR) method. The target sequence was amplified with using One–Step Invitrogen Super Script III RT-PCR System with Platinum Taq DNA Polymerase (Invitrogen Life Technologies, Carlsbad CA) using primers *pol* RT F 5’-TTC CCA TTA GTC CTA TTG AAA CTG T-3’ and RT R 5’-TCA TTG ACA GTC CAG CTA TCC TTT T-3’ in a 25 μl reaction. Cycling conditions were 45°C for 40 min and 94°C for 1 min in first-round RT-PCR, followed by 30 cycles at 94°C for 1 min,58°C for 1 s,72°C for 1min, and an extension at 72°C for 10 min. The nested PCR was performed using Super Script III Taq Platinum PCR kit using primers *pol* F1 5′-CAGAGCCAACAGCCCCACCA-3′ and *pol* F2 5’-CCATCCAAAGAAATGGAGGTTC-3′ in a 25 μl reaction and the cycling conditions were 95°C for 5 min in first-round RT-PCR, followed by 35 cycles at 94°C for 30s,58°C for 30 s,72°C 2.5 min, and an extension at 72°C for 10 min. PCR purification was done using Sure Extract Spin PCR Clean-up/Gel Extraction kit (Genex Biotech, New Delhi, India). The PCR purified product was detected and analyzed on 1% agarose gel. PCR products corresponding to the RT coding regions were sequenced. Purified product (15-45ng) was used for sequencing, using forward primer and big dye terminator ready reaction mix (ABI Big Dye Terminator version 3.0 Ready Reaction Cycle Sequencing Kit, Applied Biosystems, USA) according to the manufacturer’s instructions. For sequencing reaction, template (15-45ng), forward primer 5’-GCCTGAAAATCCATATAACACTCC-3’ (1.6µl of 10pmol concentration) of the *pol* gene, PCR buffer (1.5µl) and water (variable) was added to 2-4µl ABI Prism Big Dye (which contains DNA polymerases, dNTPs and labeled ddNTPs). 10µl of the reaction was set up for sequencing and the samples were subjected to thermo cycler GeneAmp® PCR system 9700 (Applied Biosciences, USA). The reaction mixture was brought at 4°C and then at -20°C till further use. Purification of the sequencing reaction mix, i.e., removal of the unincorporated dye, unused primer and unused dNTPs was carried out according to version used. For version V3.1; 12µl master mix I was added to 10µl sequencing reaction followed by the addition of 52µl master mix II. Contents were mixed and incubated at room temperature for 15 min. and the centrifuged at 12000 rpm for 20 min. Finally, the pellet was dissolved in 10-20µl template suppressor reagents (TSR) and the contents were denatured at 95°C for 2min. followed by immediate chilling on ice. The samples were then loaded on to the sequencing machine.

### Phylogenetic analysis

Phylogenetic trees were constructed for *pol* gene RNA sequences that included at least the first (1-260 codons) of the reverse transcriptase gene. Phylogenetic analysis was carried out with 30 worldwide reference sequences. The reference sequences used were from China (JQ658754, HG421606, KJ614011), Myanmar (N223216, JN223183, and KC913773), Nepal (JN023039 JN023035, KX430115), India (DQ838761, AY746371, EU68378), South Africa (DQ222286, EF381756), Botswana (AY829288, EF026185, AY829270), Swaziland (KY274409, EU244650, KY274425), Zimbabwe (AY998363, AY0908, AY998351), Norway (GQ398881, GQ400431), United Kingdom (AY714442), and Ethiopia (AB285760, AB285752)obtained from Los Alamos National Laboratory HIV sequence database of common HIV-1 subtype C; HIV-1 subtype K-K03455|HIVHXB2 (Human immunodeficiency virus type 1 (HXB2), complete genome; HIV1/HTLV-III/LAV reference genome) from the United States of America was used as an outgroup species.

All the sequences were imported into the Molecular Evolutionary Genetic Analysis Software version 11 (MEGA 11.0).The imported sequences were aligned in Clustal W. The MEGA 11.0 best-fit model feature was used to select the best fit model for nucleotide substitution resulting in HKY+G (Hasegawa-Kishino-Yano) with the discrete gamma categories of two. A maximum likelihood tree was constructed under the selected model. The reliability of the tree topology was estimated by performing bootstrap analysis with 100 bootstrap replicates as the maximum likelihood method of tree construction is computationally extensive.

## Result

The characteristics of HRGs with recent HIV infection are presented in table 1. Majority of the respondents were literate and did not have much travel outside their usual place of residence.

**Table 1:** Characteristics of HRGs with recent HIV infection

| **Characteristics**<br>**(N-37)** | **FSW**<br>**n=9** | **MSM**<br>**n=7** | **IDU**<br>**n=21** |
| --- | --- | --- | --- |
| **Age-group (in years)** |  |  |  |
| 18-24 | 2(22.2) | 1(14.3) | 2(9.5) |
| 25 and above | 7(77.8) | 6(85.7) | 19(90.5) |
| **Marital Status** |  |  |  |
| Never Married | 1(11.1) | 1(14.3) | 6(28.6) |
| Ever Married | 8(88.9) | 6(85.7) | 15(71.4) |
| **Literacy Status** |  |  |  |
| Illiterate | 5(55.6) | - | 5(23.8) |
| Literate | 4(44.4) | 7(100) | 16(76.2) |
| **Times Traveled** |  |  |  |
| Up to 2 | 8(88.9) | 5(71.4) | 17(81) |
| More than 2 | 1(11.1) | 2(28.6) | 4(19) |
| **Places Traveled** |  |  |  |
| Up to 2 | 7(77.8) | 3(42.9) | 13(61.9) |
| More than 2 | 2(22.2) | 4(57.1) | 8(38.1) |
| **State** |  |  |  |
| Haryana | 5(55.6) | 2(28.6) | 2(9.5) |
| Himachal Pradesh | 1(11.1) | 1(14.3) | 1(4.8) |
| Chandigarh | 1(11.1) | 1(14.3) | 4(19) |
| Punjab | 2(22.2) | 3(42.9) | 14(66.7) |

### Phylogenetic analysis

Phylogeny of isolates from FSW, MSM and IDU are presented in figure 1,2, and 3.

**Figure 1:**
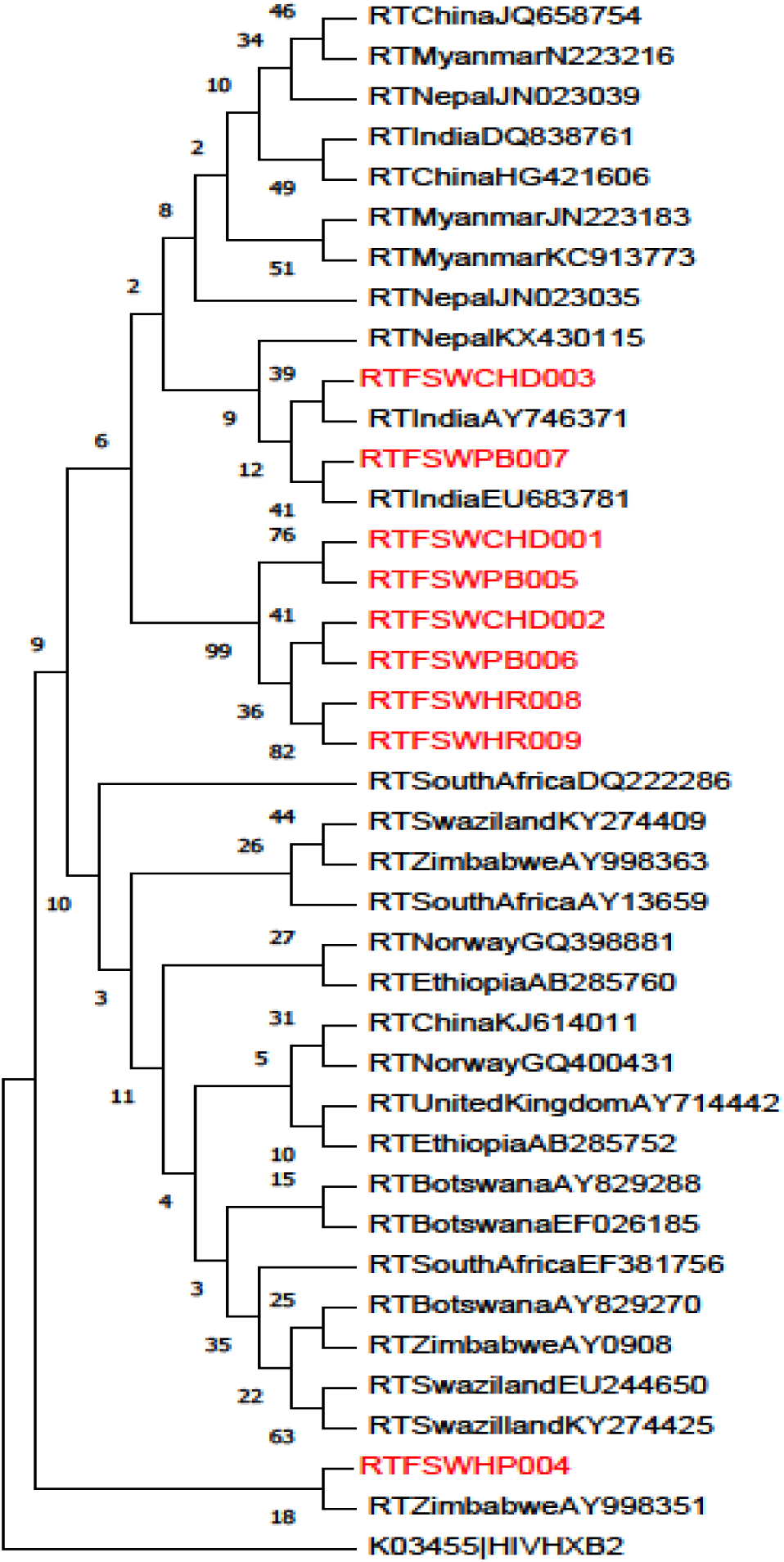
The number of base substitutions per site from between sequences are shown. Analyses were conducted using the Kimura 2-parameter model [17]. The rate variation among sites was modeled with a gamma distribution (shape parameter = 5). This analysis involved 37 nucleotide sequences. Codon positions included were 1st+2nd+3rd+Noncoding. All ambiguous positions were removed for each sequence pair (pairwise deletion option). There were a total of 536 positions in the final dataset. Evolutionary analyses were conducted in MEGA11 [18]

**Figure 2:**
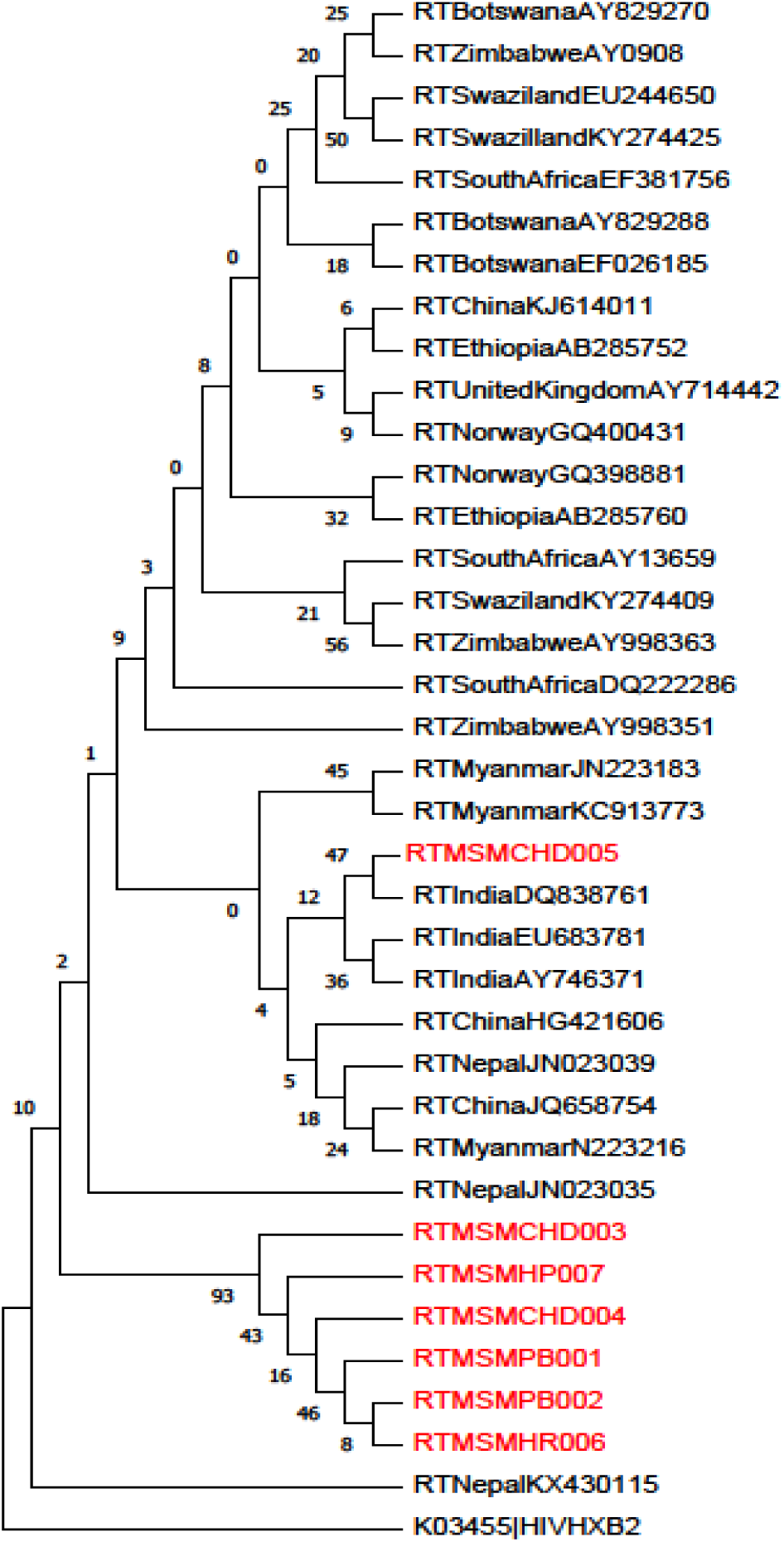
The number of base substitutions per site from between sequences are shown. Analyses were conducted using the Kimura 2-parameter model [17]. The rate variation among sites was modeled with a gamma distribution (shape parameter = 5). This analysis involved 37 nucleotide sequences. Codon positions included were 1st+2nd+3rd+Noncoding. All ambiguous positions were removed for each sequence pair (pairwise deletion option). There were a total of 537 positions in the final dataset. Evolutionary analyses were conducted in MEGA11 [18]

**Figure 3:**
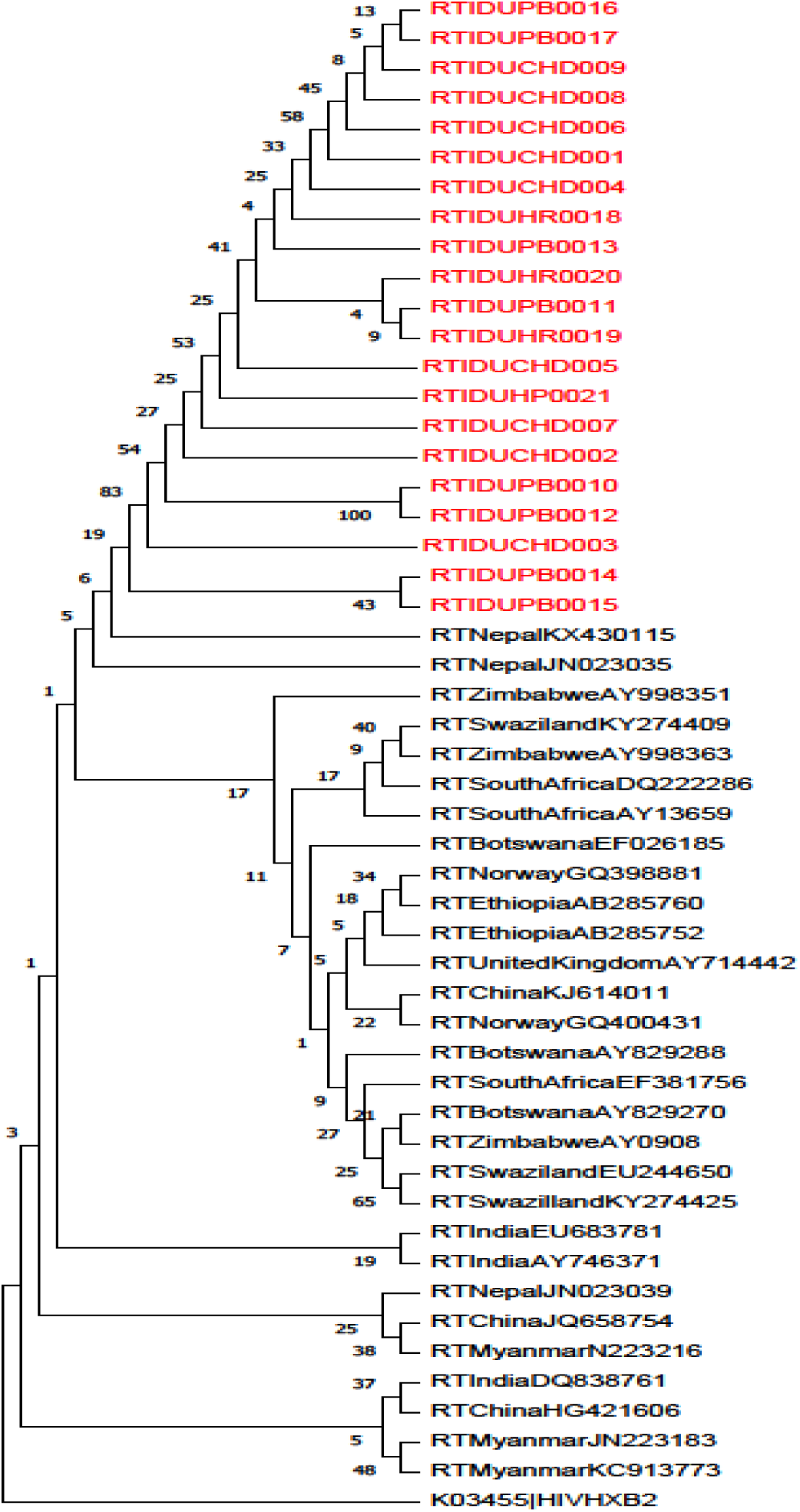
The numbers of base substitutions per site from between sequences are shown. Analyses were conducted using the Kimura 2-parameter model [17]. The rate variation among sites was modeled with a gamma distribution (shape parameter = 5). This analysis involved 51 nucleotide sequences. Codon positions included were 1st+2nd+3rd+Noncoding. All ambiguous positions were removed for each sequence pair (pairwise deletion option). There were a total of 535 positions in the final dataset. Evolutionary analyses were conducted in MEGA11 [18]

FSW: The phylogeny of FSW data clearly depicts that the study isolates RTFSWCHD003 and RTFSWPB007 cluster with and are related to the Indian reference sequences AY746371 and EU683781 and a Nepal sequence KX430115. The other six study isolates-RTFSWCHD001, RTFSWPB005 RTFSWCHD002 RTFSWPB006 RTFSWHR008 RTFSWHR009 clustered uniquely among themselves without any interlinking with other references. Another study isolate RTFSWHP004, clustered closely with Zimbabwe isolate AY998351.

MSM: The phylogeny of MSM data shows that the study isolate MSMCHD005 clades separately with the Indian references DQ838761, EU683781,and AY746371, but is also very closely related to the references from China (HG421606, JQ658754), Nepal(JN023039) and Myanmar (N223216, JN223183, KC913773). The other study isolates MSMCHD003, MSMHP007, MSMCHD004, MSMPB001, MSMPB002, and MSMHR006 are highly interrelated among themselves and form a separate unique clade altogether.

IDU: The constructed evolutionary tree of IDU data shows that all the sequences from our current study formed a monophyletic lineage i.e., sequences from India clustered together more than sequences from any other country. The study sequences showed relatedness only to the Nepal references KX430115 and JN023035. The South African, UK, Norway, China, and Myanmar references are grouped into an altogether separate clade.

## Discussion

The phylogenomic analyses showed that majority of the isolates were clustered with Indian subtype C. In present study 3 phylogenetic trees were made separately for each group HRG (FSW, MSM and IDU). In the three phylogenetic trees, a total of nine transmission clusters were observed. FSW isolates were closely related to each other and one isolate of FSW show close networks with Zimbabwe, all MSMs cluster separately showing their high interrelatedness among themselves and one MSM study isolate forms a close cluster with only Indian reference sequences from Pune. IDU isolates showed closed networks.

The behavioral patterns of these groups supported our networking patterns because the trend of HIV epidemic in India is exclusively concentrated among various high risk groups. While FSW have been the pioneer of the epidemic since first few cases of HIV were observed in South India. Migration is an important risk factor in HIV transmission [6,7]. Extensive migration can expand HIV epidemics geographically by bridging high-risk sexual networks in multiple locations [8]. Moreover, mobile FSWs are more vulnerable to HIV due to inconsistent access to preventive program services and less able to adapt and negotiate safer sex practices given their lack of influence with those controlling the sex work environment [9]. They may have clients who are less familiar to them and therefore might be unable to negotiate condom use [7]

In present study the genetic interpretation revealed that MSM have relatively closed networks with few open connections. This confirms the existence of a potential network among MSM living in this sub-continent. Previous epidemiological studies emphasized that MSM are more vulnerable to HIV infection. These studies also revealed that factors like high number of male partners, unprotected anal intercourse (especially receptive), and high prevalence of sexually transmitted diseases (STD) along with mobility among MSMs are some of the major hurdles associated with this high risk group to contain the infection [10,11]. Thus, various levels viz individual-level, network-level and community-level risk turn helpful for a high HIV transmission rates in MSM populations, because larger networks directly translate into more opportunities for exposure to different sexual practices and HIV-positive potential partners [12].The transmission relationship is compound, and migration among MSMs would lead to bi-directional spread of HIV isolates [13].In present study, majority of MSMs hailed from Punjab Haryana, Himachal Pradesh and Chandigarh and 99% of MSMs had migrated to other districts. Previous studies have also concluded that the high mobility of MSM contributes to the nationwide and a more complex spread of HIV epidemic in MSM [14,15].However, in the present study mostly MSM networks were found to be within the region.There is an indication in the present study that IDUs were interlinked together within India only. None of the samples from IDU group showed interlinking with other country’s samples.

Present study reported that most infections acquired were intra-national and that the HIV-1 subtype C is still largely predominant circulating among IDUs and HRGs in general in Indian subcontinent. A study conducted by Neogi *et al* (2012) concluded that phylogenomic analyses of the three viral genes (*gag, env* and *pol)* demonstrated that a vast majority of all Indian HIV-1C strains in the analysis grouped together in the principle Indian clades. This shows that most infections are obtained inside the nation, but detailed regional transmission patterns couldn’t be discerned. An extraordinary admixing was seen among the northern, western, focal and southern Indian strains. The *pol* phylogenetic tree recognized a little clade for the most part comprising of the strains from western, focal and north-eastern districts of the nation, demonstrating development of strains among these locales [16].

## Conclusions

The transmission patterns among high risk groups can reveal critical information relevant to the control of HIV epidemic. The methods used in the present study can help in timely contact tracing, risk reduction, early treatment with antiretroviral agents which would be a progressive to interrupt transmission.

## Data Availability

Yes available upon reasonable request to the authors

## Acknowledgments

The authors would like to acknowledge study participants who gave blood sample and personal history, without who study would not have been possible.

## Author contributions

RK, PVML, SKA and CKC contributed towards the conception and design of this study. RK, PVML, VS, AS and CKC contributed to the study implementation; CKC participated in collection of the biological samples, analysis of biological samples and PS constructed the phylogenetic trees. CKC and PS wrote the manuscript. All authors read and approved the final manuscript. RK is the guarantor of the paper.

## References

1. Klovdahl AS. Social networks and the spread of infectious diseases: the AIDS example. Social Science & Medicine. 1985;21(11):1203–16.

2. Smith DM, May SJ, Tweeten S, Drumright L, Pocold ME, Pond SLK, et al. A Public Health Model for the molecular surveillance of HIV transmission in San Diego, California. AIDS. 2009;23(2):225–32.

3. Weniger BG, Takebe Y, Ou CY, Yamazaki S. The molecular epidemiology of HIV in Asia. AIDS. 2001; 15(4):545.

4. Grabowski MK, Redd AD. Molecular tools for studying HIV transmission in sexual networks. CurrOpin HIV and AIDS. 2014;9(2):126–33.

5. Chauhan CK, Lakshmi PVM, Sagar V, Sharma A, Arora SK, Kumar R. Immunological markers for identifying recent HIV infection in North-West India. Indian J Med Res.2020;153:227–233.

6. Quinn TC, Overbaugh J. HIV/AIDS in women: an expanding epidemic. Science.2005; 308 (5728):1582–3.

7. Webber G. The Impact of Migration on HIV Prevention for Women: Constructing a Conceptual Framework. Health Care for Women International. 2007; 28(8):712–30.

8. Blanchard JF, Neil JO, Ramesh BM, Bhattacharjee P, Orchard T, Moses S. Understanding the social and cultural contexts of female sex workers in Karnataka, India: implications for prevention of HIV infection. J Infect Dis. 2005;191(1): S139–46.

9. Quinn TC. Population migration and the spread of types 1 and 2 human immunodeficiency viruses. Proceedings of the National Academy of Sciences of the United States of America. 1994;91(7):2407–14.

10. Wang L, Wang L, Norris JL, Li DM, Guo W, Ding ZW, et al. HIV prevalence and influencing factors analysis of sentinel surveillance among men who have sex with men in China, 2003–2011. Chin Med J. 2012;125(11):1857–61.

11. Lau JT, Lin C, Hao C, Wu X, Gu J. Public health challenges of the emerging HIV epidemic among men who have sex with men in China. Public Health. 2011;125(5):260–5.

12. Sullivan P, Carballo-Dieguez A, Coates T, Goodreau SM, McGowan Sandors EJ, et al. Successes and challenges of HIV prevention in men who have sex with men. Lancet. 2012;380(9839):388–99.

13. Chen M, Ma Y, Su Y, Yang L, Zang R, Yang C, et al. HIV-1 Genetic Characteristics and Transmitted Drug Resistance among Men Who Have Sex with Men in Kunming, China. PLoS ONE. 2014;9(1):e87033.

14. Wong FY, Huang ZJ, He N, Smith BD, Ding Y, Fu C, et al. HIV risks among gay- and non-gay-identified migrant money boys in Shanghai, China. AIDS Care. 2008;20(2):p170–180

15. Wang B, Li X, Stanton B, Liu Y, Jiang S. Socio-demographic and behavioral correlates for HIV and syphilis infections among migrant men who have sex with men in Beijing, China. AIDS Care. 2013;25(2):249–257.

16. Neogi U, Bonetell I, Shet A, Costa AD, Gupta S, Diwan V, et al. Molecular epidemiology of HIV 1 subtypes in India. PLoS ONE. 2012;7(6):e39819

17. Kimura M. A simple method for estimating evolutionary rate of base substitutions through comparative studies of nucleotide sequences. Journal of Molecular Evolution.1980;16:111–120.

18. Tamura K., Stecher G., and Kumar S. MEGA 11: Molecular Evolutionary Genetics Analysis Version 11. Molecular Biology and Evolution. 2021. https://doi.org/10.1093/molbev/msab120.

